# Telehealth Utilization and Patient Experiences: The Role of Social Determinants of Health Among Individuals with Hypertension and Diabetes

**DOI:** 10.1101/2024.08.01.24311392

**Authors:** Haoxin Chen, Will Simmons, Malak Abu Hashish, Jiancheng Ye

## Abstract

**Objective:** To evaluate the utilization patterns, effectiveness, and patient satisfaction of telehealth services among individuals with hypertension and/or diabetes, and to investigate the influence of social determinants of health (SDOH) on telehealth access and utilization in this population.

**Methods:** We conducted a cross-sectional analysis using data from the 2022 Health Information National Trends Survey (HINTS 6) by the National Cancer Institute. The study sample included 3,009 respondents with self-reported diabetes, hypertension, or both conditions. Telehealth usage was assessed through 14 survey questions, and participant characteristics were analyzed using sociodemographic, baseline health, and SDOH data.

**Results:** Of the 6,252 HINTS 6 survey respondents, 3,009 met the inclusion criteria. Significant sociodemographic differences were observed across the diabetes and/or hypertension groups. No significant differences were found in telehealth usage among the groups, with 43.9% of respondents utilizing telehealth in the past year. Common reasons for telehealth use included provider recommendation, convenience, and infection avoidance. Social determinants of health, such as food insecurity and transportation issues, were more prevalent among individuals with both conditions, though no significant differences in telehealth experiences were noted across groups.

**Conclusion:** Telehealth shows potential for managing chronic conditions like hypertension and diabetes, demonstrating substantial adoption and universal accessibility. However, disparities influenced by SDOH highlight the need for targeted interventions to ensure equitable access. Addressing privacy concerns, leveraging healthcare providers’ recommendations, and tackling SDOH barriers are crucial for fostering wider telehealth adoption and improving outcomes. Future research should focus on the long-term impacts of telehealth and further investigate SDOH factors to develop tailored interventions that enhance engagement and equitable access across diverse patient populations.

## INTRODUCTION

Chronic diseases such as hypertension and diabetes are prevalent health concerns that significantly impact individuals’ quality of life and pose substantial challenges to healthcare systems worldwide [1]. Hypertension, a leading cause of cardiovascular diseases, and diabetes, a major contributor to morbidity and mortality, often co-occur, exacerbating health complications and increasing the burden on affected individuals. Managing these conditions effectively requires comprehensive and continuous care, which can be complicated by various social determinants of health (SDOH) [2].

SDOH, including socioeconomic status, education, neighborhood and physical environment, employment, and social support networks, play a crucial role in shaping health outcomes. Individuals with lower socioeconomic status, limited education, and inadequate access to healthcare resources are more likely to experience poor health outcomes and have higher rates of chronic conditions. These determinants influence not only the prevalence of hypertension and diabetes but also the ability of individuals to manage these conditions effectively. Addressing SDOH is therefore essential for improving health outcomes and reducing health disparities among populations with chronic diseases [3]. In addition to understanding the impact of SDOH and the utilization of telehealth, it is crucial to consider patients’ willingness to share information about their SDOH with healthcare providers. Sharing such information can significantly enhance care coordination and allow for more personalized and comprehensive treatment plans [4]. However, privacy concerns and the sensitivity of the information can influence patients’ willingness to disclose these details. Understanding the factors that affect patients’ willingness to share SDOH information can help in designing interventions that address privacy concerns and foster trust between patients and healthcare providers [5].

Telehealth has emerged as a promising solution to enhance access to healthcare services, particularly for individuals managing multiple chronic conditions [6]. The COVID-19 pandemic accelerated the adoption of telehealth, demonstrating its potential to provide convenient and effective healthcare remotely [7]. Telehealth services offer several benefits, including increased accessibility, reduced travel time, and the ability to maintain regular monitoring and management of chronic conditions [8, 9]. However, the utilization and effectiveness of telehealth can be influenced by various factors, including technology access, digital literacy, and individual preferences.

This study aims to elucidate telehealth experiences and challenges in managing hypertension and diabetes by comparing statistical outcomes among telehealth participants with these conditions [9, 10]. Furthermore, we aim to examine the influence of SDOH on telehealth access and utilization in this population [11]. By investigating factors like income, education, housing stability, and transportation access, the study aims to identify disparities in telehealth usage based on socioeconomic factors. Understanding how SDOH impact telehealth engagement can inform targeted interventions to promote equitable access and reduce healthcare disparities among vulnerable populations [12].

## METHODS

### Study Design

We conducted a cross-sectional analysis of data from the 2022 Health Information National Trends Survey (HINTS 6) of the National Cancer Institute. HINTS surveys are conducted annually among noninstitutionalized US civilians ≥ 18 years and contain nationally representative data on awareness and use of health-related information [13]. This study used HINTS 6 data to evaluate differences in telehealth use and related SDOH among participants with diabetes, hypertension, or both conditions. The HINTS 6 sample comprises 6,252 respondents, with surveys conducted from March 7, 2022 to November 8, 2022. We excluded participants who did not report diabetes or hypertension status, as well as who reporting having neither condition.

The Health Information National Trends Survey (HINTS) provides data on the American public’s health-related behaviors, perceptions, and access to information. HINTS 6 sheds light on the utilization patterns, effectiveness, and patient satisfaction of telehealth services among individuals with hypertension and/or diabetes.[14, 15].

### Measures

We divided the sample into three groups: individuals with diabetes only, those with hypertension only, and those with both diabetes and hypertension. Participants’ diabetes and hypertension status was determined using two yes-or-no questions: “Has a doctor or other health professional ever told you that you had (1) diabetes or high blood sugar; (2) high blood pressure or hypertension?” We characterized the sample using participants’ self-reported sociodemographic data, including age, sex, occupational status, marital status, level of education, race, ethnicity, sexual orientation, household income, rural-urban residence status, and U.S. geographic region. Additionally, we summarized participants’ health data, encompassing self-rated health, heart and lung conditions, depression/anxiety, and cigarette use, as well as social determinants of health such as food insecurity, housing, transportation, and their comfort with sharing related information with healthcare providers.

### Statistical Analysis

HINTS utilized a complex sampling design, which we accounted for by incorporating sample weights using the Taylor Series (linear approximation) method. All analyses were performed on weighted data in R version 4.3.1, with the srvyr package used for complex sample weighting and the survey package used for survey-appropriate statistical tests. Summary tables were created using the tbl_svysummary function in the gtsummary package.

To assess differences in participant sociodemographic, baseline health, social determinants of health, and telehealth use by diabetes/hypertension category, we used the Wilcoxon rank-sum test adapted for complex survey samples [16] or the chi-squared test with Rao & Scott’s second-order correction for continuous and categorical variables, respectively.[17]

All reported p-values and confidence intervals (CI) were two-sided and adjusted for multiple comparisons using the Benjamini-Hochberg procedure. The type I error rate was set to 0.05.

### Study Approval

We solely used publicly available HINTS data for this study.

## RESULTS

Of the total HINTS 6 sample comprising 6,252 individuals, 30 respondents (weighted 0.5%) were missing diabetes status, 22 (weighted 0.4%) were missing hypertension status, and 222 (weighted 3.6%) were missing both and were therefore excluded. Additionally, 2,969 respondents (weighted 47.5%) reported having neither condition. Thus, the final sample comprised 3,009 individuals with either diabetes, hypertension, or both.

**Table 1** presents sociodemographic characteristics across diabetes-only, hypertension-only, and diabetes-hypertension groups. Variables such as sex, tobacco use, urban or rural residence, and census regions did not significantly differ among the groups, suggesting minimal impact on hypertension or diabetes status. Significant differences were observed in age (p<0.001); participants with both hypertension and diabetes had a mean (SD) age of 64.0 (16.3), the highest among the three groups. Regarding occupational status, respondents with only diabetes had a higher proportion of employment, while those with both conditions were more likely to be retired (p<0.001). For marital status, respondents with only diabetes had a higher proportion in divorced status, but fewer in widowed (p=0.007). In terms of education level, respondents with both diabetes and hypertension had more individuals with less than 8 years of education and fewer people with a college degree or higher (p=0.001). For race (p<0.001) and ethnicity (p<0.001), more participants with hypertension were White, and more with both conditions were Black or African American. Participants with conditions had relatively lower household incomes (p<0.001). Overall, individuals with both conditions tended to be older, retired, non-Hispanic Black or African American, and had lower education and household incomes.

**Table 1.** Characteristics of participants.

| **Variables** | **Hypertension only (N=1714)** | **Diabetes only (N=317)** | **Hypertension+ Diabetes (N=978)** | **P value** |
| --- | --- | --- | --- | --- |
| **Age, mean (SD), years** | 61.5 (17.1) | 57.2 (16.6) | 64.0 (16.3) | <0.001 |
| **Sex, n (%)** |  |  |  | 0.476 |
| Male | 688 (41.1) | 117 (38.9) | 404 (42.7) |  |
| Female | 987 (58.9) | 184 (61.1) | 543 (57.3) |  |
| **Occupational status, n (%)** |  |  |  | <0.001 |
| Employed | 655 (39.0) | 135 (44.9) | 253 (26.5) |  |
| Unemployed for 1 year or more | 36 (2.1) | 15 (5.0) | 16 (1.7) |  |
| Unemployed for less than 1 year | 23 (1.4) | 3 (1.0) | 9 (0.9) |  |
| Homemaker | 39 (2.3) | 11 (3.7) | 30 (3.1) |  |
| Student | 10 (0.6) | 2 (0.7) | 3 (0.3) |  |
| Retired | 679 (40.5) | 81 (26.9) | 429 (45.0) |  |
| Disabled | 95 (5.7) | 27 (9.0) | 98 (10.3) |  |
| Other | 11 (0.7) | 1 (0.3) | 7 (0.7) |  |
| Multiple Occupation statuses selected | 130 (7.7) | 26 (8.6) | 108 (11.3) |  |
| **Marital status, n (%)** |  |  |  | 0.007 |
| Married | 749 (44.8) | 129 (43.3) | 407 (42.8) |  |
| Living as married or with a romantic partner | 85 (5.1) | 17 (5.7) | 31 (3.3) |  |
| Divorced | 311 (18.6) | 68 (22.8) | 177 (18.6) |  |
| Widowed | 251 (15.0) | 26 (8.7) | 164 (17.3) |  |
| Separated | 37 (2.2) | 12 (4.0) | 31 (3.3) |  |
| Single | 240 (14.3) | 46 (15.4) | 140 (14.7) |  |
| **Education, n (%)** |  |  |  | 0.001 |
| Less than 8 years | 27 (1.6) | 5 (1.7) | 32 (3.4) |  |
| 8-11 years | 92 (5.5) | 16 (5.3) | 57 (6.0) |  |
| 12 years or completed high school | 311 (18.6) | 62 (20.6) | 228 (24.0) |  |
| Post high school other than college | 149 (8.9) | 22 (7.3) | 83 (8.7) |  |
| Some college | 374 (22.3) | 62 (20.6) | 223 (23.4) |  |
| College graduate | 425 (25.4) | 86 (28.6) | 204 (21.5) |  |
| Postgraduate | 296 (17.7) | 48 (15.9) | 124 (13.0) |  |
| **Ethnicity, n (%)** |  |  |  | <0.001 |
| Non-Hispanic White | 981 (62.2) | 126 (45.5) | 415 (47.1) |  |
| Non-Hispanic Black or African American | 296 (18.8) | 45 (16.2) | 217 (24.6) |  |
| Hispanic | 196 (12.4) | 80 (28.9) | 178 (20.2) |  |
| Non-Hispanic Asian | 56 (3.6) | 14 (5.1) | 41 (4.6) |  |
| Non-Hispanic Other | 47 (3.0) | 12 (4.3) | 31 (3.5) |  |
| **Race, n (%)** |  |  |  | <0.001 |
| White only | 1162 (71.3) | 193 (67.7) | 543 (59.8) |  |
| Black only | 336 (20.6) | 55 (19.3) | 257 (28.3) |  |
| American Indian or Alaska Native only | 16 (1.0) | 4 (1.4) | 11 (1.2) |  |
| Multiple races | 47 (2.9) | 14 (4.9) | 37 (4.1) |  |
| Asian Indian only | 14 (0.9) | 1 (0.4) | 9 (1.0) |  |
| Chinese only | 13 (0.8) | 7 (2.5) | 8 (0.9) |  |
| Filipino only | 12 (0.7) | 4 (1.4) | 16 (1.8) |  |
| Vietnamese only | 5 (0.3) | 2 (0.7) | 6 (0.7) |  |
| Other Asian only | 18 (1.1) | 1 (0.4) | 7 (0.8) |  |
| Other Pacific Islander only | 6 (0.4) | 4 (1.4) | 14 (1.5) |  |
| **Sexual orientation, n (%)** |  |  |  | 0.219 |
| Heterosexual | 1543 (94.4) | 268 (93.7) | 850 (93.1) |  |
| Homosexual | 40 (2.4) | 7 (2.4) | 17 (1.9) |  |
| Bisexual | 29 (1.8) | 4 (1.4) | 23 (2.5) |  |
| Else | 22 (1.3) | 7 (2.4) | 23 (2.5) |  |
| **Income, n (%)** |  |  |  | <0.001 |
| \$0 to \$9,999 | 114 (7.3) | 23 (8.0) | 100 (11.3) | |
| \$10,000 to \$14,999 | 85 (5.5) | 23 (8.0) | 82 (9.2) | |
| \$15,000 to \$19,999 | 84 (5.4) | 11 (3.8) | 47 (5.3) | |
| \$20,000 to \$34,999 | 209 (13.5) | 47 (16.4) | 135 (15.2) | |
| \$35,000 to \$49,999 | 209 (13.5) | 40 (14.0) | 149 (16.8) | |
| \$50,000 to \$74,999 | 270 (17.4) | 51 (17.8) | 141 (15.9) | |
| \$75,000 to \$99,999 | 208 (13.4) | 27 (9.4) | 99 (11.2) | |
| \$100,000 to \$199,999 | 278 (17.9) | 44 (15.4) | 99 (11.2) | |
| \$200,000 or more | 96 (6.2) | 20 (7.0) | 35 (3.9) | |
| **Body Mass Index (BMI), mean (SD), kg/m<sup>2</sup>** | 28.9 (9.0) | 29.3 (9.3) | 30.2 (10.1) | 0.002 |
| **General health, n (%)** |  |  |  | <0.001 |
| Excellent | 88 (5.2) | 14 (4.4) | 27 (2.8) |  |
| Very good | 526 (30.9) | 101 (32.1) | 176 (18.2) |  |
| Good | 752 (44.2) | 113 (35.9) | 422 (43.6) |  |
| Fair | 288 (16.9) | 76 (24.1) | 279 (28.9) |  |
| Poor | 48 (2.8) | 11 (3.5) | 63 (6.5) |  |
| **Heart condition, n (%)** | 240 (14.0) | 28 (8.9) | 234 (24.0) | <0.001 |
| **Chronic lung disease, n (%)** | 282 (16.5) | 53 (16.8) | 204 (20.9) | 0.014 |
| **Mental health problem, n (%)** | 448 (26.2) | 98 (31.0) | 298 (30.5) | 0.028 |
| **Tobacco user, n (%)** |  |  |  | 0.998 |
| Current (Everyday) | 142 (8.5) | 27 (8.8) | 82 (8.6) |  |
| Former (Some days) | 60 (3.6) | 10 (3.3) | 33 (3.5) |  |
| Never (Not at all) | 1475 (88.0) | 269 (87.9) | 841 (88.0) |  |
| **Rural/Urban, n (%)** |  |  |  | 0.111 |
| Metropolitan | 1435 (83.7) | 278 (87.7) | 832 (85.1) |  |
| Micropolitan | 155 (9.0) | 24 (7.6) | 72 (7.4) |  |
| Small town | 91 (5.3) | 9 (2.8) | 44 (4.5) |  |
| Rural | 33 (1.9) | 6 (1.9) | 30 (3.1) |  |
| **Census, n (%)** |  |  |  | 0.289 |
| Northeast | 238 (13.9) | 38 (12.0) | 127 (13.0) |  |
| Midwest | 289 (16.9) | 48 (15.1) | 151 (15.4) |  |
| South | 831 (48.5) | 147 (46.4) | 496 (50.7) |  |
| West | 356 (20.8) | 84 (26.5) | 204 (20.9) |  |

Health characteristics also varied across the three respondent groups. Significant differences were noted in BMI (p=0.002), self-rated health (p<0.001), presence of heart conditions (p<0.001), chronic lung disease (p=0.014), and mental health problems (p=0.028). Participants with both hypertension and diabetes had higher BMIs than the other groups, while a greater proportion of them rated their general health as good or fair, and had heart disease, chronic lung disease, and mental health problems compared to the other two groups.

**Table 2** and **Figures 1** illustrate telehealth use across the three groups. In general, approximately 43% of respondents utilized telehealth in the past year, citing reasons such as provider recommendation, convenience, and infection avoidance. We found significantly different rates of receiving a telehealth visit over the past year among participants with hypertension only, diabetes only, and both conditions (p=0.001). Specifically, the breakdown of hypertension participants receiving telehealth is as follows: 298 (17.7%) received telehealth care by video, 248 (14.7%) by phone calls, 175 (10.4%) by both video and phone calls, and 961 (57.1%) not engaging in telehealth visits. For diabetes participants: 66 (21.4%) received telehealth care by video, 56 (18.1%) by phone calls, 28 (9.1%) by both video and phone calls, and 159 (51.5%) with no telehealth visit. For participants with both conditions: 160 (16.8%) by video, 204 (21.4%) by phone calls, 93 (9.8%) by both ways, and 496 (52.0%) with no telehealth visit.

**Figure 1.**
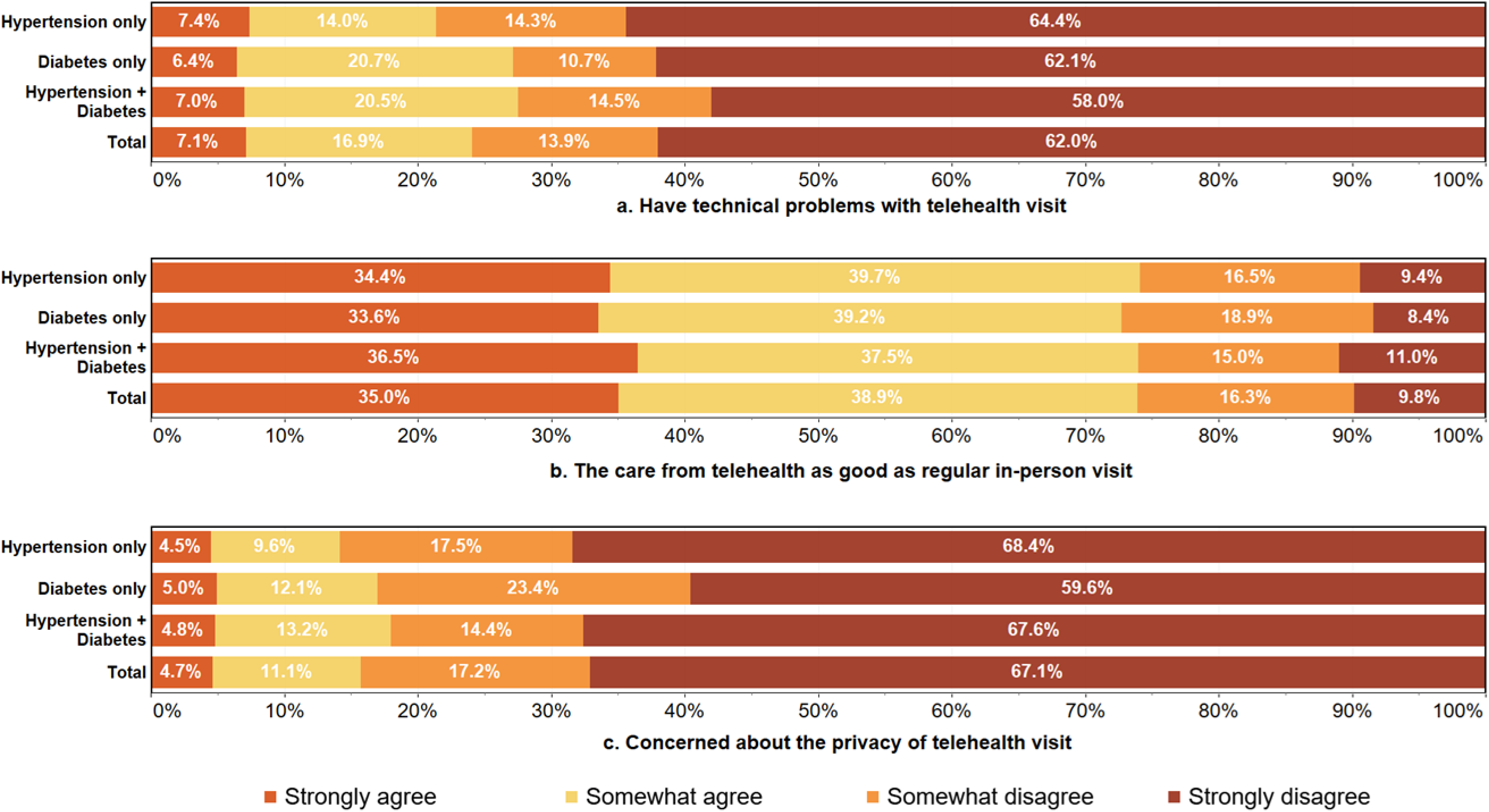
Experiences with using telehealth across the three groups.

**Table 2.** Characteristics of telehealth utilization and patient experiences.

| Variables | Hypertension<br>only (N=1714) | Diabetes<br>only (N=317) | Hypertension+<br>Diabetes<br>(N=978) | P<br>value |
| --- | --- | --- | --- | --- |
| **Received telehealth care, n (%)** |  |  |  | 0.001 |
| Yes, by video | 298 (17.7) | 66 (21.4) | 160 (16.8) |  |
| Yes, by phone call (no video) | 248 (14.7) | 56 (18.1) | 204 (21.4) |  |
| Yes, some video some phone call | 175 (10.4) | 28 (9.1) | 93 (9.8) |  |
| No telehealth visits | 961 (57.1) | 159 (51.5) | 496 (52.0) |  |
| **Offered telehealth option, n (%)** |  |  |  | 0.034 |
| Yes | 161 (17.4) | 20 (13.2) | 101 (21.5) |  |
| No | 636 (68.8) | 114 (75.0) | 323 (68.9) |  |
| Did not try | 127 (13.7) | 18 (11.8) | 45 (9.6) |  |
| **Primary reason for telehealth, n (%)** |  |  |  | 0.002 |
| Annual visit | 161 (23.5) | 29 (20.0) | 100 (23.8) |  |
| Minor illness | 148 (21.6) | 23 (15.9) | 53 (12.6) |  |
| Manage chronic health | 206 (30.1) | 50 (34.5) | 171 (40.7) |  |
| Medical emergency | 13 (1.9) | 3 (2.1) | 9 (2.1) |  |
| Mental health | 68 (9.9) | 20 (13.8) | 28 (6.7) |  |
| Other | 88 (12.9) | 20 (13.8) | 59 (14.0) |  |
| **Regarding telehealth visits, how much do you agree or disagree n (%)** |  |  |  |  |
| **a. I had technical problems with telehealth** |  |  |  | 0.102 |
| Strongly agree | 49 (7.4) | 9 (6.4) | 28 (7.0) |  |
| Somewhat agree | 93 (14.0) | 29 (20.7) | 82 (20.5) |  |
| Somewhat disagree | 95 (14.3) | 15 (10.7) | 58 (14.5) |  |
| Strongly disagree | 428 (64.4) | 87 (62.1) | 232 (58.0) |  |
| **b. Telehealth was as good care as in-person visits** |  |  |  | 0.839 |
| Strongly agree | 234 (34.4) | 48 (33.6) | 153 (36.5) |  |
| Somewhat agree | 270 (39.7) | 56 (39.2) | 157 (37.5) |  |
| Somewhat disagree | 112 (16.5) | 27 (18.9) | 63 (15.0) |  |
| Strongly disagree | 64 (9.4) | 12 (8.4) | 46 (11.0) |  |
| **c. I had privacy concerns about telehealth** |  |  |  | 0.157 |
| Strongly agree | 30 (4.5) | 7 (5.0) | 19 (4.8) |  |
| Somewhat agree | 64 (9.6) | 17 (12.1) | 52 (13.2) |  |
| Somewhat disagree | 116 (17.5) | 33 (23.4) | 57 (14.4) |  |
| Strongly disagree | 454 (68.4) | 84 (59.6) | 267 (67.6) |  |

Regarding being offered a telehealth visit option, feedback from the three groups was also significantly diverse (p=0.034). Only 161 (17.4%) participants with hypertension, 20 (13.2%) with diabetes and 101 (21.5%) with both conditions were suggested the option of telehealth when they schedule medical care, which indicates that telehealth does not seem to be widespread in current days yet. For the primary reasons of recent telehealth visit, the statistic was significantly different among the groups (p=0.002). For participants with hypertension, 206 (30.1%) were for managing chronic health, followed by 161 (23.5%) for annual visits, 148 (21.6%) for minor illness, 88 (12.9%) for other reasons, 68 (9.9%) for mental health, and 13 (1.9%) for medical emergency. For participants with diabetes, 50 (34.5%) were for managing chronic health, followed by 29 (20.0%) for annual visits, 23 (15.9%) for minor illness, 20 (13.8%) for mental health and other reasons, and 3 (2.1%) for medical emergency. For participants with two conditions, 171 (40.7%) were for managing chronic health, 100 (23.8%) for annual visits, 59 (14.0%) for other reasons, 53 (12.6%) for minor illness, 28 (6.7%) for mental health, and 9 (2.1%) for medical emergency. Apparently, most patients utilize telehealth for their long-term health conditions and chronic diseases, instead of emergencies requiring urgent and in-person treatment. **(Table 2)**

We also noted that most individuals who experienced telehealth visit had little technical obstacles, with overall 62.0% strongly disagreed with having troubles and 13.9% somewhat disagreed. 35.0% of them strongly agreed and 38.9% somewhat agreed that the care from telehealth was as good as in-person visit. 67.1% of them did not have severe privacy concerns.**(Figure 1)** However, no statistically significant difference among the groups on their thoughts and experiences with telehealth visit were found, particularly, having technical problems with telehealth (p=0.102), regarding telehealth as good as regular visit (p=0.839), and having privacy concerns with telehealth (p=0.157). **(Table 2)** Similarly, we performed a descriptive analysis of the reasons for not selecting and selecting a telehealth visit. From the result, many individuals did not specify their answers, and there were no statistically significant differences among the groups. With respect to the reasons for not choosing a telehealth visit, over 90% of people in general reflected that they preferred an in-person visit, whereas for nearly 80% of people, privacy concerns were not their main consideration. Additionally, about 27% of people thought telehealth would be difficult to use.

**Table 3** demonstrates the participants’ reasons for choosing telehealth, over 70% of people agreed that they chose to have a telehealth visit because of the recommendation by their healthcare provider. Only about 25% of people were asking for advice on whether in-person care is needed. About half of them admitted that they wanted to avoid potential infection in hospital, and 57% of them believed that it was more convenient than going to the doctor, saving travelling and waiting time. However, these statistics are not significantly different among the three groups. They held different opinions only on the reason that they could include family or other caregivers in the appointment (p=0.022), with 25.9% of participants with both hypertension and diabetes considering more in this factor.

**Table 3:**
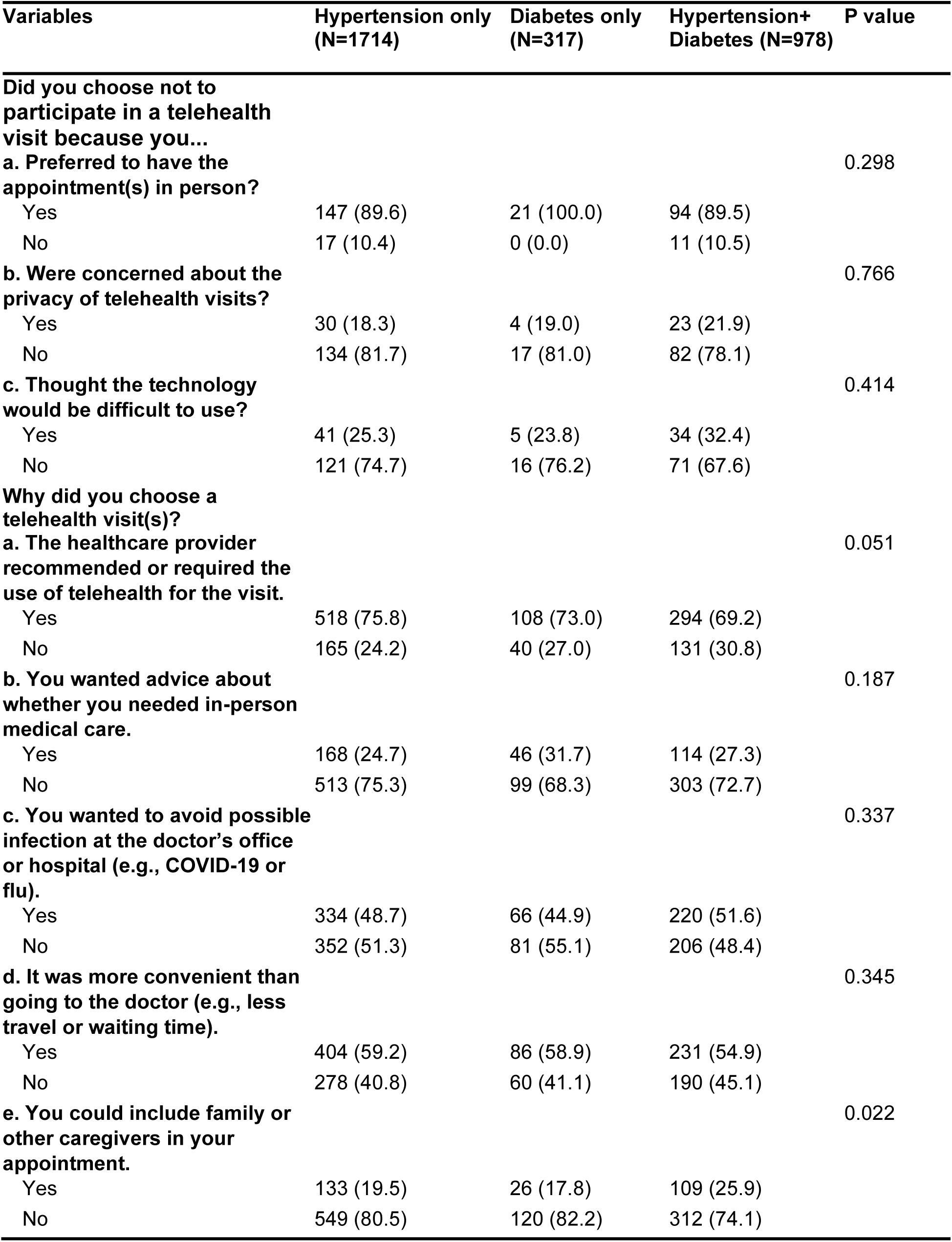
Reasons for not choosing or choosing a telehealth visits.

**Figure 2** and **Figure 3** depict the social determinants of health across groups. Concerns such as food insecurity, affordability of balanced meals, worries about relocation, and transportation issues were prevalent across all groups. **Figure 2** shows that 86.5% of people in total did not need to compromise on meals, 84.2% could afford balanced nutrition, and 87.5% did not face challenges in accessing reliable transportation for their daily needs, although the situation of participants with both hypertension and diabetes was slightly worse than the other two groups. 89.2% of people in general experienced concerns about relocation, while those with only diabetes tended to worry more. Overall, people with only hypertension have the best situation among the groups, while people with both hypertension and diabetes faced more difficulties.

**Figure 2.**
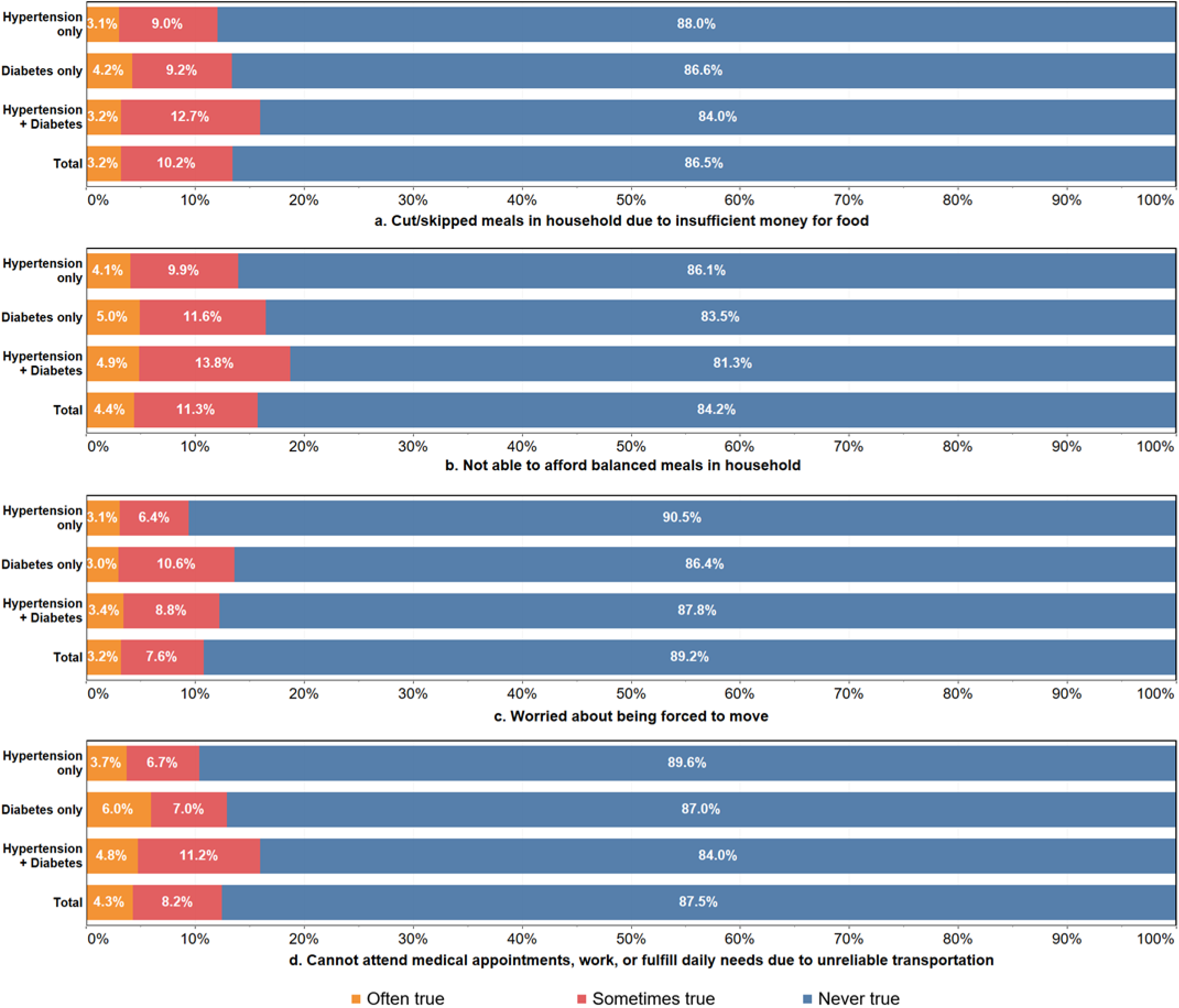
Characteristics of social determinants of health across the three groups.

**Figure 3.**
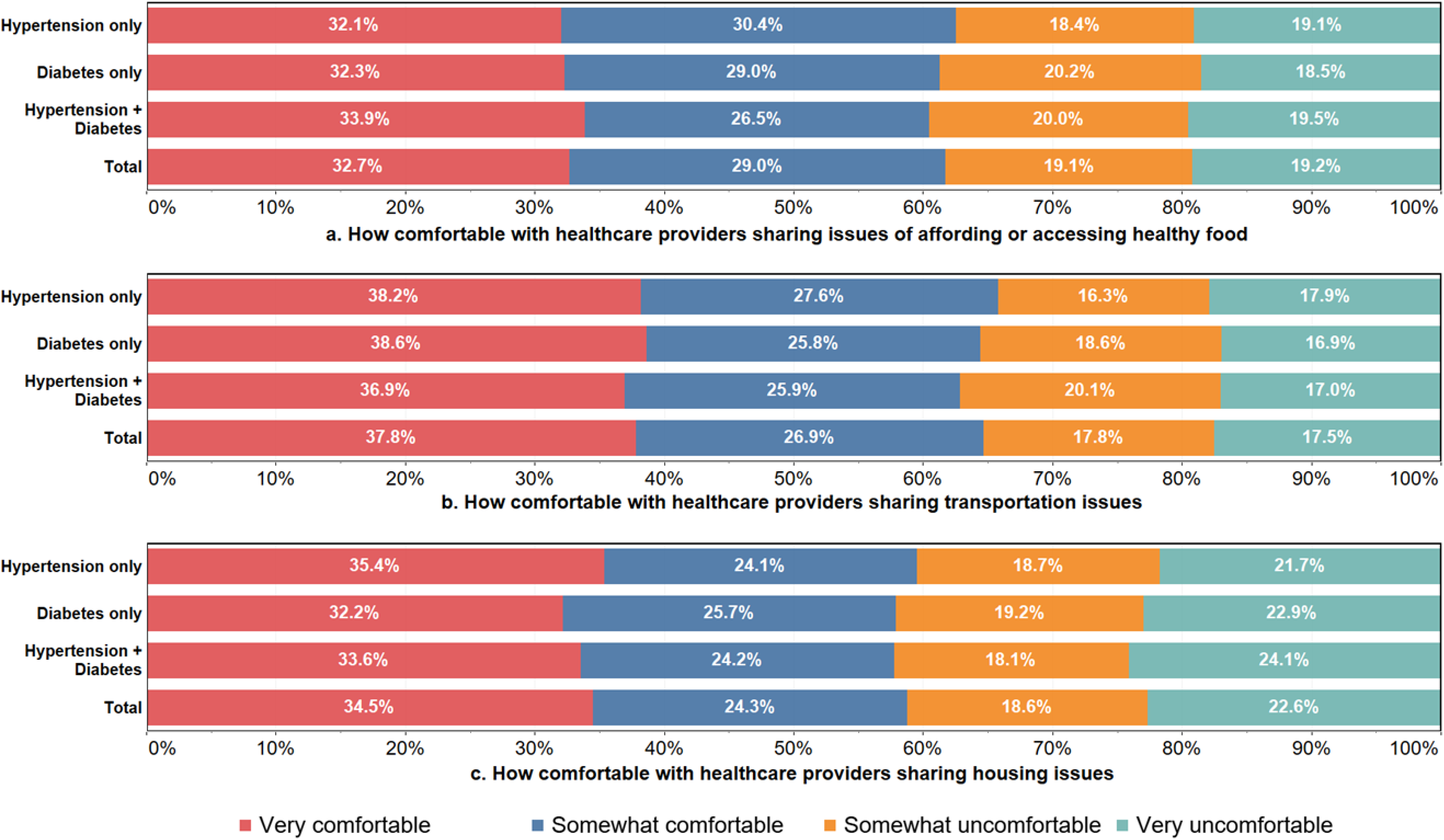
Willingness to share social determinants of health information with health care providers across the three groups.

**Figure 3** illustrates the Opinions on whether participants would be willing to allow health care providers to share each other their related information on food, transportation and housing for treatment purposes, which were relatively evenly distributed and had little difference among three groups. Concerning the food issues, 32.7% of people were very supportive and 29.0% felt comfortable about it, while the rest felt a bit offensive. For the transportation issue, people were slightly more comfortable sharing the information, with 37.8% felt completely comfortable and 26.9% comfortable. However, 22.6% of people felt strongly uncomfortable and 18.6% felt uncomfortable if their housing issues were shared. In general, people tended to be more concerned about the privacy of their housing conditions.

## DISCUSSION

The findings from our analysis provide valuable insights into the utilization of telehealth among individuals managing hypertension and/or diabetes. By examining how factors like age, health status, and social determinants of health (SDOH) influence their experiences with telehealth, we uncovered important patterns. Notably, we found no significant differences in telehealth experiences across groups with varying health conditions, including hypertension, diabetes, or both. This underscores telehealth’s potential as a universally accessible and acceptable mode of health care delivery.

### Privacy and equity in telehealth adoption

Concerns about privacy were noted among a minority of participants, highlighting the importance of addressing privacy and security issues in telehealth platforms to ensure patient trust [18, 19]. To address these concerns, healthcare organizations and telehealth providers should implement robust security measures, such as end-to-end encryption, secure data storage, and strict access controls [20].

Conducting telehealth services should also be transparent about the privacy policies and procedures, clearly communicating to patients how their personal information will be protected and how it will be used [21]. Providing patients with the ability to control the sharing of their data and offering options for anonymous consultations can further enhance their sense of privacy and security [22]. By prioritizing patient privacy and addressing any concerns in this area, telehealth providers can foster greater trust and confidence among users, ultimately encouraging more widespread and effective utilization of these valuable healthcare services [18, 23].

Additionally, the reasons for choosing telehealth varied among respondents, with convenience and infection avoidance being primary factors. Interestingly, healthcare provider recommendation emerged as a significant influencer of telehealth utilization, suggesting the pivotal role of healthcare professionals in promoting and facilitating telehealth adoption among patients [24, 25]. While convenience and healthcare provider recommendations are significant drivers of telehealth adoption, efforts should also be directed towards addressing systemic barriers and disparities in access [26]. Initiatives to improve digital literacy, expand broadband infrastructure, and enhance reimbursement policies for telehealth services can help mitigate these barriers and ensure equitable access to care [12, 27, 28].

Participants’ willingness to share information on food, transportation, and housing with healthcare providers varied, with the most concern expressed about sharing housing information. This reluctance to share personal information could be attributed to privacy concerns and the sensitive nature of housing issues. Healthcare providers should ensure that data sharing is conducted transparently and securely to build trust with patients and encourage information sharing that can enhance care coordination and support [29]. However, our study found no statistically significant differences among the groups regarding their willingness to share information about food and transportation. This suggests a general apprehension towards disclosing SDOH information, which could hinder the effectiveness of interventions targeting these social factors. Strategies to address these concerns and educate patients on the benefits of sharing SDOH information are necessary for improving care coordination and outcomes.[30]

### Patients managing comorbidities

The study’s findings highlight a significant uptake of telehealth services, with 43.9% of respondents utilizing them over the past year [14]. This emphasizes the growing importance of telehealth in healthcare, especially during the COVID-19 pandemic, where remote care has become crucial due to safety measures [31].

The effective management of multiple chronic conditions through telehealth presents a promising avenue for enhancing patient outcomes. Our results indicate that telehealth services are frequently used for managing long-term health conditions and chronic diseases rather than for urgent care needs. This is particularly important for individuals with both hypertension and diabetes, who require continuous monitoring and management of their conditions.[32] Telehealth stands to transform hypertension and diabetes management by facilitating regular monitoring and remote consultations, reducing the need for in-person visits, and easing travel burdens, especially for rural or underserved populations [23, 33]. Improved accessibility through telehealth can enhance treatment adherence and overall disease management [34]. However, accessing and adopting telehealth services may be more challenging for patients managing multiple chronic conditions [35]. The difference in telehealth use among these groups stems from various factors. Individuals managing a single chronic condition, like hypertension or diabetes, may find telehealth convenient for tasks such as monitoring health metrics and managing medications [11, 12, 36]. They face fewer competing healthcare needs and logistical hurdles, making it easier to integrate telehealth into their care routine [37].

On the other hand, patients with both diabetes and hypertension face a heavier care burden, requiring more frequent interactions with healthcare providers and specialized treatments [38, 39]. The complexity of their healthcare needs may hinder the effective incorporation of telehealth into their care plan, with challenges like accessing necessary technology and coordinating virtual appointments with multiple providers.[40] Additionally, those contending with multiple chronic conditions may encounter barriers related to digital literacy or socioeconomic status, such as limited access to reliable internet or devices [41, 42]. These disparities in technology access disproportionately affect certain patient groups, exacerbating telehealth utilization inequalities [43].

To tackle these disparities, healthcare systems and policymakers should implement targeted interventions to support patients managing multiple chronic conditions in accessing and utilizing telehealth services effectively [12, 44]. This may involve providing digital literacy training,[45] offering loaner devices or internet connectivity solutions, and collaborating with community organizations to address social determinants of health impacting telehealth access [46, 47]. By addressing these systemic barriers, healthcare providers can ensure equitable access to the convenience and accessibility of telehealth services for all patients, regardless of their health status or socioeconomic circumstances [12, 31].

### Addressing social determinants of health challenges

Our study explored how social determinants of health (SDOH) influence access to and use of telehealth services. We found that individuals managing both diabetes and hypertension often face more challenges, such as food insecurity and transportation limitations, compared to other demographic groups [14]. This highlights the significance of considering broader social and economic factors that can affect access to healthcare services, including telehealth. Factors like income, education, housing stability, transportation, and food security all play a significant role in a person’s ability to use telehealth effectively [11, 31].

By addressing these underlying social determinants of health, healthcare providers and policymakers can take significant strides towards ensuring equitable access to telehealth services for all individuals, regardless of their socio-economic circumstances [12, 44]. This may involve implementing targeted interventions such as providing subsidies for internet access, offering transportation assistance,[48] or partnering with community organizations to address food insecurity. Addressing these disparities is crucial to ensuring equitable access to telehealth services and reducing healthcare inequities [49].

### Limitations

This study has several limitations. The cross-sectional design limits the ability to establish causality between the observed characteristics and health outcomes. The data in this study relies on participants’ self-reported responses, which could introduce biases. Recall bias may occur if participants do not accurately remember or report their behaviors and experiences. Social desirability bias is also a concern, as participants might provide responses they think are more socially acceptable, rather than reflecting their true experiences [50]. Additionally, the study sample might not fully represent the broader population, potentially limiting how widely we can apply our findings [51–53].

Moving forward, it is important for research to investigate the long-term effects of using telehealth on healthcare outcomes for people managing hypertension and diabetes[54]. Additionally, further investigation into the social determinants of health factors driving the observed disparities is warranted [55]. This includes developing tailored interventions to enhance telehealth engagement and improve outcomes across diverse patient demographics. [12].

## CONCLUSION

This study highlights the substantial adoption of telehealth among individuals managing hypertension and diabetes, underscoring its potential to revolutionize chronic disease management. Our findings reveal no significant differences in telehealth experiences across different health condition groups, emphasizing its universal accessibility. However, disparities influenced by SDOH underscore the need for targeted interventions to ensure equitable access. Addressing privacy concerns and leveraging healthcare provider recommendations are crucial for fostering wider telehealth adoption. Future research should focus on the long-term impacts of telehealth and further investigate SDOH factors to develop tailored interventions that enhance engagement and improve outcomes across diverse patient populations. By prioritizing these areas, we can maximize telehealth’s benefits, ensuring it serves as a transformative tool in healthcare delivery.

## CONTRIBUTION STATEMENT

JY designed the study, contributed to the data analyses, and the writing of the manuscript. HC, WS, and MAH contributed to data analyses and the writing of the manuscript. All authors read and approved the final version of the manuscript.

## DATA AVAILABILITY STATEMENT

All data are available at the National Cancer Institute Website. The relevant code and analyses are available at: https://github.com/ConstanzaChen/Hyper_telehealth.git

